# OPPORTUNISTIC ASSESSMENT OF CARDIOVASCULAR RISK USING AI-DERIVED STRUCTURAL AORTIC AND CARDIAC PHENOTYPES FROM NON-CONTRAST CHEST COMPUTED TOMOGRAPHY

**DOI:** 10.1101/2025.01.28.25321302

**Authors:** Daniel W. Oo, Audra Sturniolo, Matthias Jung, Marcel Langenbach, Borek Foldyna, Douglas P. Kiel, Hugo JWL Aerts, Pradeep Natarajan, Michael T. Lu, Vineet K. Raghu

## Abstract

**Background:** Primary prevention of cardiovascular disease relies on accurate risk assessment using scores such as the Pooled Cohort Equations (PCE) and PREVENT. However, necessary input variables for these scores are often unavailable in the electronic health record (EHR), and information from routinely collected data (e.g., non-contrast chest CT) may further improve performance. Here, we test whether a risk prediction model based on structural features of the heart and aorta from chest CT has added value to existing clinical algorithms for predicting major adverse cardiovascular events (MACE).

**Methods:** We developed a LASSO model to predict fatal MACE over 12 years of follow-up using structural radiomics features describing cardiac chamber and aorta segmentations from 13,437 lung cancer screening chest CTs from the National Lung Screening Trial. We compared this radiomics model to the PCE and PREVENT scores in an external testing set of 4,303 individuals who had a chest CT at a Mass General Brigham site and had no history of diabetes, prior MACE, or statin treatment. Discrimination for incident MACE was assessed using the concordance index. We used a binary threshold to determine MACE rates in patients who were statin-eligible or ineligible by the PCE/PREVENT scores (≥7.5% risk) or the radiomics score (≥5.0% risk). Results were stratified by whether all variables were available to calculate the PCE or PREVENT scores.

**Results:** In the external testing set (n = 4,303; mean age 61.5 ± 9.3 years; 47.1% male), 8.0% had incident MACE over a median 5.1 years of follow-up. The radiomics risk score significantly improved discrimination beyond the PCE (c-index 0.653 vs. 0.567, p < 0.001) and performed similarly in individuals who were missing inputs. Those statin-eligible by both the radiomics and PCE scores had a 2.6-fold higher incidence of MACE than those eligible by the PCE score alone (29.5 [20.5, 39.1] vs. 11.2 [8.0, 14.4] events per 1,000 person-years among PCE-eligible individuals). In patients missing inputs, incident MACE rates were 1.8-fold higher in those statin-eligible by the radiomics score than those statin-ineligible (29.5 [21.9, 37.6] vs. 16.7 [14.3, 19.0] events per 1000 person-years). Similar results were found when comparing to the PREVENT score. Left ventricular volume and short axis length were most predictive of myocardial infarction, while left atrial sphericity and surface-to-volume ratio were most predictive of stroke.

**Conclusions:** Based on a single chest CT, a cardiac shape-based risk prediction model predicted cardiovascular events beyond clinical algorithms and demonstrated similar performance in patients who were missing inputs to standard cardiovascular risk calculators. Patients at high-risk by the radiomics score may benefit from intensified primary prevention (e.g., statin prescription).

## INTRODUCTION

Cardiovascular diseases are the leading cause of death worldwide (1), in part due to limitations in accurately identifying high-risk individuals. The American College of Cardiology and American Heart Association guidelines recommend using a risk calculator such as the Pooled Cohort Equations (PCE) for adults aged 40-75 years without diabetes mellitus and with low-density lipoprotein cholesterol between 70-190 mg/dL (2). The PCE is a widely-used regression model that estimates 10-year atherosclerotic cardiovascular disease risk based on cardiovascular risk factors (e.g., demographics, cholesterol, blood pressure). The recently proposed PREVENT equations from the American Heart Association (3) incorporate additional risk factors (BMI, estimated glomerular filtration rate) to improve miscalibration of the PCE in modern populations (4) and predict 10-year and 30-year outcomes, including heart failure. However, these calculators are still imprecise (5,6) and not all patients have the necessary variables in the electronic health record (EHR) to calculate PCE or PREVENT risk–additional readily available information may better personalize risk estimates to guide preventive care, such as statin initiation.

Routine imaging, such as non-contrast enhanced chest computed tomography (CT), is a promising way to non-invasively assess cardiovascular health to refine risk estimates. Non-contrast chest CTs are a common radiologic imaging exam, often ordered for lung cancer screening, evaluation of cough and chest pain, and treatment planning for surgical or radiation therapy (7). Recent advances in deep learning enable accurate, automated segmentation of anatomical structures (8) and measurement of coronary artery calcium and adipose tissue depots from CT (9,10). Structural phenotypes of cardiovascular structures, such as the shape and size of the cardiac chambers and aorta, could provide additional information to predict cardiovascular risk. Though the association of cardiac and aortic size with cardiovascular risk is well-known (9,12), a composite risk score incorporating these phenotypes has not been developed.

Here, we investigate whether aortic and cardiac chamber structure and size measured from chest CT are associated with cardiovascular events beyond prevalent risk factors. We use a freely available open source deep learning model called TotalSegmentator (8) to identify the cardiac chambers and the aorta on non-contrast enhanced chest CT and conduct a “radiomics” analysis (13) to extract quantitative features (e.g., sphericity and volume) describing each region. We then develop a composite risk score based on these measures and test whether this score improves risk estimation beyond the PCE and PREVENT scores. Our study provides insight into the role of cardiac structure in cardiovascular disease risk and examines the clinical utility of opportunistic assessment of structural heart and aorta phenotypes from CT.

## METHODS

### Study Populations

A study overview is presented in Figure 1. In all study datasets, we identified patients with no history of myocardial infarction or stroke who had a non-contrast, non-ECG-gated chest computed tomography scan. We limited our cohort to this population as these individuals are recommended for 10-year risk assessment by current American College of Cardiology and American Heart Association guidelines for primary cardiovascular prevention (2).

**Figure 1:**
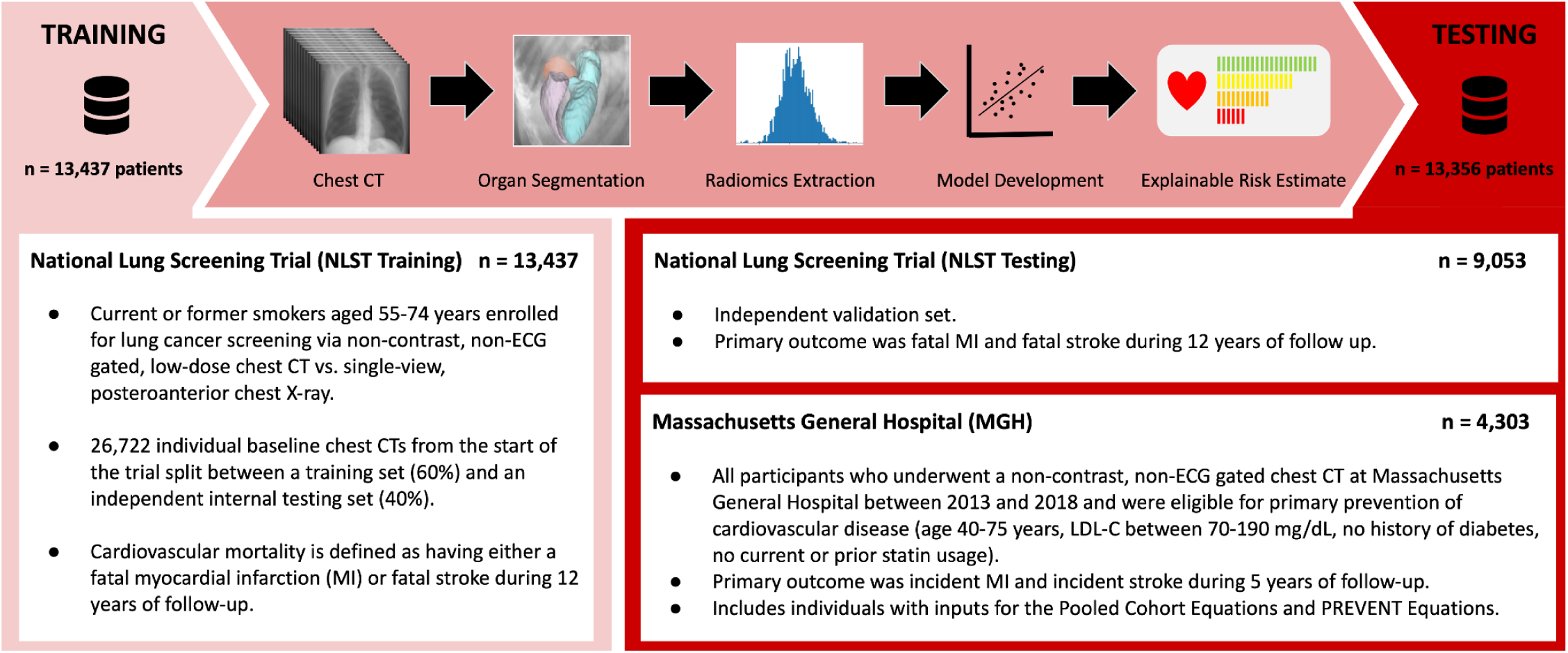
Overview of radiomics workflow.

The model was developed using chest CTs from the CT arm of the National Lung Screening Trial (NLST) (14), a multicenter imaging trial of 53,454 current or former heavy smokers aged 55 to 74 years, with >30 pack-year smoking history and no signs, symptoms, or history of lung cancer. In NLST, subjects from 33 medical institutions were randomized to either non-contrast, non-ECG-gated low-dose chest CT (26,722 participants) or a single-view posteroanterior chest X-ray between 2002 and 2007. All-cause mortality was measured over a median of 12.3 years of follow-up. The trial was approved by the institutional review board at each medical center. Details about cohort selection are shown in Figure 2. Following cohort selection and CT quality control, 22,490 individual CTs were split into a training set of 13,437 participants and a testing set of 9,053 participants.

**Figure 2:**
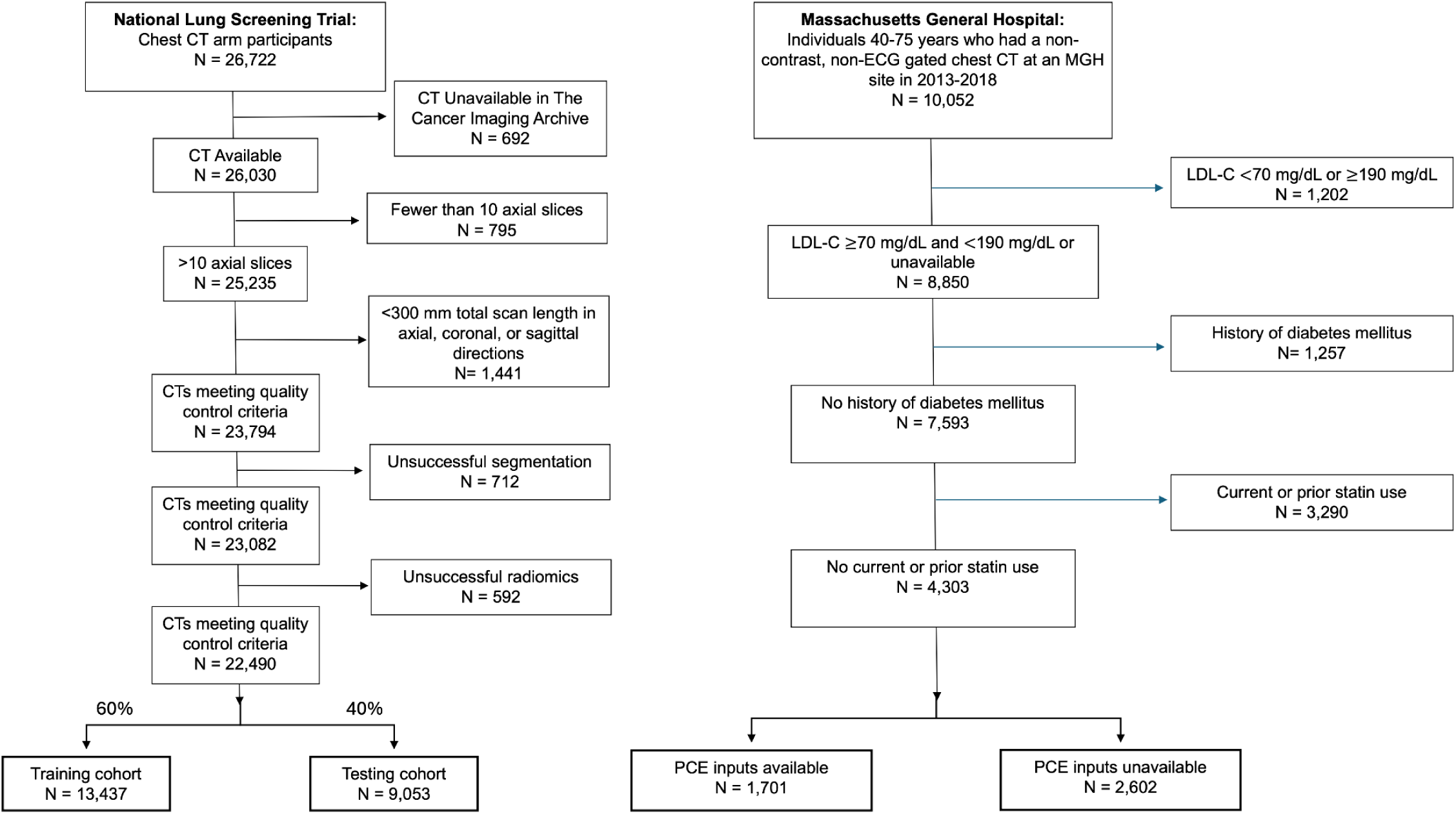
Consort diagram.

**Figure 3:**
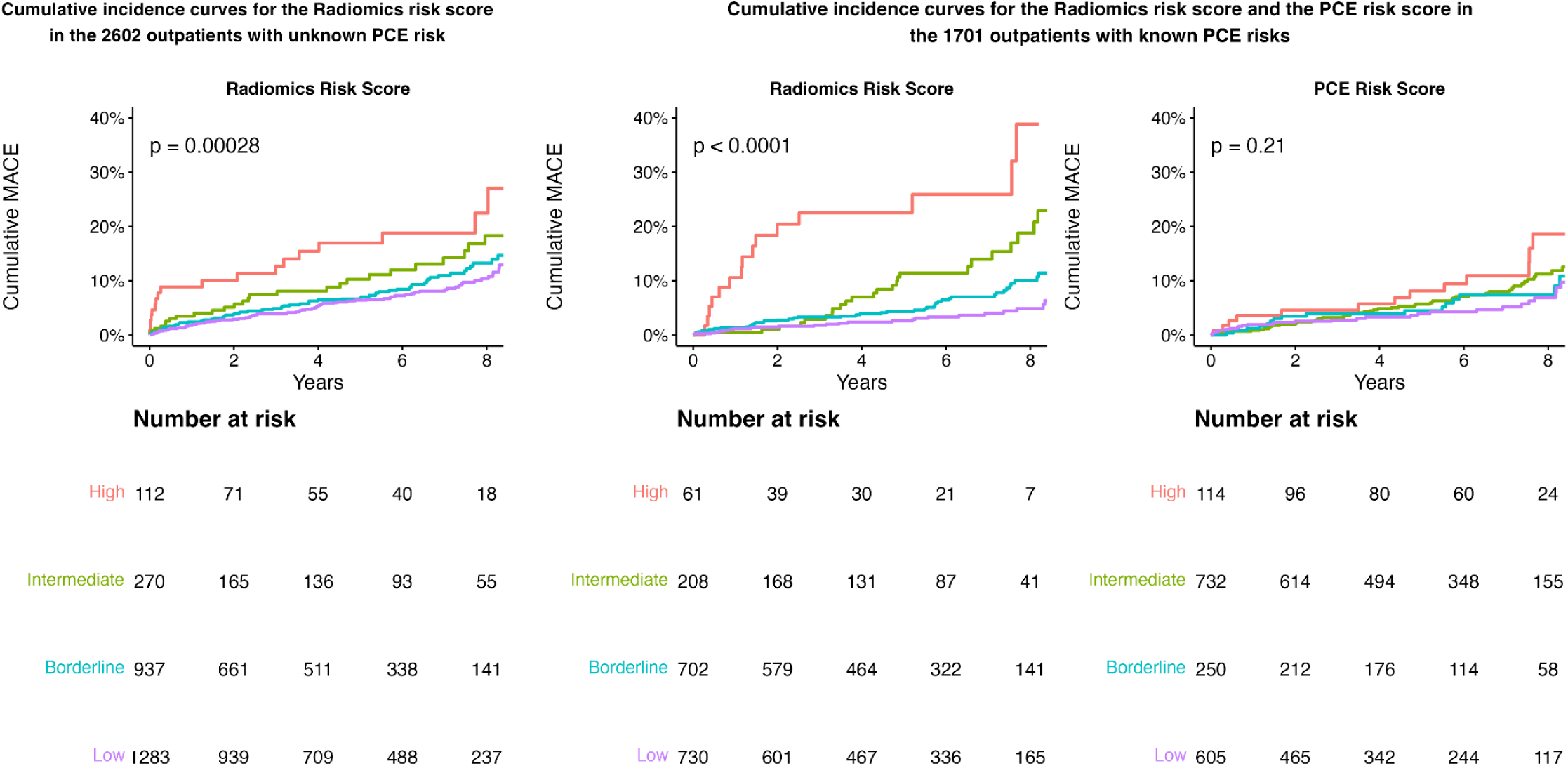
Cumulative incidence curves for the radiomics risk score in those without inputs to calculate the PCE score (left) and curves for the radiomics risk score (center) and PCE score (right) in those with inputs available to calculate the PCE risk score.

We then externally validated the model using images from patients who had a non-contrast, non-ECG-gated chest CT at a Massachusetts General Hospital (MGH) site between 2013-2018. CT quality control used the same criterion as the NLST dataset. We included only patients with no prior history of myocardial infarction or stroke, were aged 40-75 years, had LDL-C between 70-190 mg/dL, were non-diabetic, and had no statin usage (i.e. patients for whom risk estimation is indicated by current ACC/AHA guidelines; N=4,303). Incident myocardial infarction and incident stroke were measured over a median of 5.1 years of follow-up.

### Radiomics Feature Extraction

Radiomics features were extracted from each chest CT scan using the following pipeline. First, CT quality control was conducted by removing all CTs with less than 10 axial slices. Following successful conversion from DICOM to NIFTI format (15), CTs with a slice thickness of ≤3 mm and a total scan length in the axial, coronal, and sagittal axes of ≥300 mm were retained. For individuals with multiple CTs meeting these criteria, only the CT series with the most slices was retained.

Following quality control, the cardiac chambers and aorta were segmented from CTs using TotalSegmentator 1.5.7 (8), Torchvision 0.14.1, and PyTorch 1.13.1 (16). TotalSegmentator is an open-source deep learning model that segments 104 anatomic structures from CTs with high accuracy (average DICE Similarity Coefficient ∼ 0.94 across structures in independent testing). The model has an nnU-Net architecture and was trained on CTs from 1,082 individuals from University Hospital Basel in Basel, Switzerland (8). Using TotalSegmentator, the right atrium, right ventricle, left atrium, left ventricle, and aorta were segmented from each CT. Subsequently, fourteen shape-based radiomics features were extracted from each segmented region with PyRadiomics 3.1.0 (13), generating 70 features in total per individual (Supplementary Table 2).

### Outcomes

The primary outcome for model validation in NLST was fatal major adverse cardiovascular events (MACE) over a median 12.3 years of follow-up. MACE was defined in NLST using ICD-9 and ICD-10 cause of death codes for cardiovascular complications, including diseases of the arteries, arterioles and capillaries, hypertensive diseases, ischemic heart diseases, heart failure, cerebrovascular diseases, and other cardiovascular diseases (Supplementary Table 1). The primary outcome for MGH external validation was MACE defined as incident myocardial infarction or incident stroke over a median 5.1 years of follow-up. Non-acute strokes, pulmonary heart disease, and chronic ischemic heart disease were excluded from this definition.

### Risk Scores

We compared the radiomics score to the PCE and PREVENT risk scores in the Massachusetts General Hospital testing set. Both scores were computed with the PooledCohorts package (v0.0.2) in R (17). The PCE risk score included age, sex, race, history of diabetes, history of smoking, systolic blood pressure, total cholesterol, high-density lipoprotein cholesterol levels, and hypertension treatment. The PREVENT risk score included BMI and estimated glomerular filtration rate in addition to the PCE variables, minus race. For both risk scores, age, sex, and self-reported race were extracted from the electronic medical record. History of diabetes was based on the presence of corresponding ICD codes (Supplementary Table 1). We used total cholesterol, HDL cholesterol, and systolic blood pressure measured within two years of the CT. If an individual had multiple measurements within two years, we took the median of the three measurements closest to the date of the CT.

### Statistical Analysis

#### Bivariate Survival Analysis

We conducted an association analysis in the NLST training set to assess the relationship between time to 12-year fatal MACE and individual radiomics features of the heart chambers and aorta. Hazard ratios were computed by fitting a separate Cox proportional hazards model for each feature with the R survival package (v3.6) (18) to predict fatal myocardial infarction, fatal stroke, and composite MACE. We report hazard ratios both unadjusted and adjusted for age, sex, and BMI per 1 SD change in each radiomics feature.

#### Radiomics Model Development

A least absolute shrinkage and selection operator (LASSO) logistic regression model was trained to predict fatal 12-year MACE (19). The inputs to the LASSO model were 70 normalized heart and aortic shape features from each of the 13,437 chest CTs in the NLST training set. Normalization of radiomics features was performed using StandardScaler in scikit-learn (20) to have a mean of 0 and a standard deviation of 1. We used the glmnet package (v4.1.8) in R for LASSO model development (21). The L1 penalty parameter λ for the LASSO model was chosen by minimizing the mean cross-validation error observed during an internal 10-fold cross-validation.

We compared the LASSO radiomics model with the clinically-used PCE and PREVENT risk scores. Since cholesterol and blood pressure were not measured as part of NLST, we compared the radiomics model to a baseline regression model trained to predict 12-year fatal MACE using age, sex, race, BMI, pack-years, current smoking, history of diabetes, history of hypertension, and history of cancer (Supplementary Table 10). In the external MGH testing set, we compared the radiomics score to the PCE and PREVENT risk scores using data extracted from the EMR.

#### Stability of Feature Selection

Stability analysis was conducted to assess the robustness of LASSO feature selection. The LASSO model was trained 1000 times on bootstrapped samples of the NLST training set of 13,437 individual chest CTs to predict fatal 12-year MACE. For each iteration, we recorded which features were selected by the LASSO model (i.e., had nonzero coefficients). Stable features were defined as having nonzero coefficients in greater than 90% of training iterations. In subanalysis, we assessed whether selected features were consistent when training separate models to predict fatal myocardial infarction and fatal stroke.

#### Stability Across CT Reconstructions

A test-retest analysis was conducted to assess the stability of each radiomics feature across multiple reconstructions from the same study that met quality control criteria. In total, 8,056 individuals had two reconstructions of baseline CTs which were used to compute the intraclass correlation coefficient (ICC) (22) and the concordance correlation coefficient (CCC) (23) for each radiomics feature. ICC was computed with the IRR package in R (v0.84.1) as a two-way agreement model. CCC was computed with the epiR package in R (v2.0.76). ICC and CCC values can be found in Supplementary Table 4.

#### Discrimination and Calibration

Model discrimination of the radiomics, PCE, and PREVENT risk scores was assessed using Harrell’s c-statistic (24) in the R Survival package (v3.6) (18). Model calibration of the radiomics score in the NLST testing set was assessed via calibration plots (Supplementary Figure 4). Calibration plots were constructed using the ggplot2 package (v3.5.1) in R (25).

#### Decision Curve Analysis

Decision curve analysis is a technique used to assess the net benefit of intervention across a range of risk thresholds for different risk prediction models. At a given threshold, net benefit is calculated by weighing the benefits and harms suggested by the threshold. For example, if a provider is willing to initiate statins for patients with >2% risk, this implies that the provider considers the harm of initiating statins in 98 patients who will not go on to have a MACE equal to the benefits of preventing MACE in 2 patients. Thus, the net benefit calculation at a 2% threshold considers 1 true positive equal to 49 false positives. We use this analysis technique to calculate the net benefit of the radiomics score, the PCE risk score, and baseline strategies to treat all patients and treat no patients across a range of possible thresholds. Decision curve analysis was conducted via the R Stats Package (v4.4.1).

#### Ordinal Risk Stratification and Statin Eligibility

Finally, we assessed the association between ordinal risk groups and incident MACE in the MGH testing dataset. PCE and PREVENT risk scores were divided into categories via standard thresholds (low, <5%; borderline, ≥5% to <7.5%; intermediate, ≥7.5% to <20%; high, ≥20%) (2). We stratified the radiomics score into ordinal groups using the following thresholds: low (<3.0%), borderline (3.0% to <5.0%), intermediate (5.0% to <7.5%), high (≥7.5%). These thresholds were selected such that the radiomics risk group had a similar number of participants as the analogous PCE risk group. We assessed the association of ordinal risk groups with incident MACE using Cox proportional hazards analysis, and by reporting incident MACE per 1000 person-years across ordinal risk categories. We repeated this analysis using a binary “statin-eligibility” threshold of PCE/PREVENT risk score ≥7.5% (2) or radiomics score ≥5.0%.

## RESULTS

### Cohort Characteristics

In the NLST training cohort of 13,437 individuals, 91.1% (N=12,200) were White, 59.2% (N=7,956) were male, and the mean age was 61.4 years (SD, 5.0) (Table 1). 2.4% (N=323) of patients died from myocardial infarction and 0.6% (N=83) of patients died from stroke over a median follow-up of 12.3 years. The internal, independent NLST testing cohort consisted of 9,053 individuals. The cohort was 90.5% (N=8,204) non-Hispanic white and 59.0% (N=5,327) male with a mean age of 61.4 years (SD, 5.1). In this testing set, 2.5% (N=230) of patients died from myocardial infarction and 0.7% (N=59) died from stroke over a median follow-up of 12.3 years.

**Table 1:**
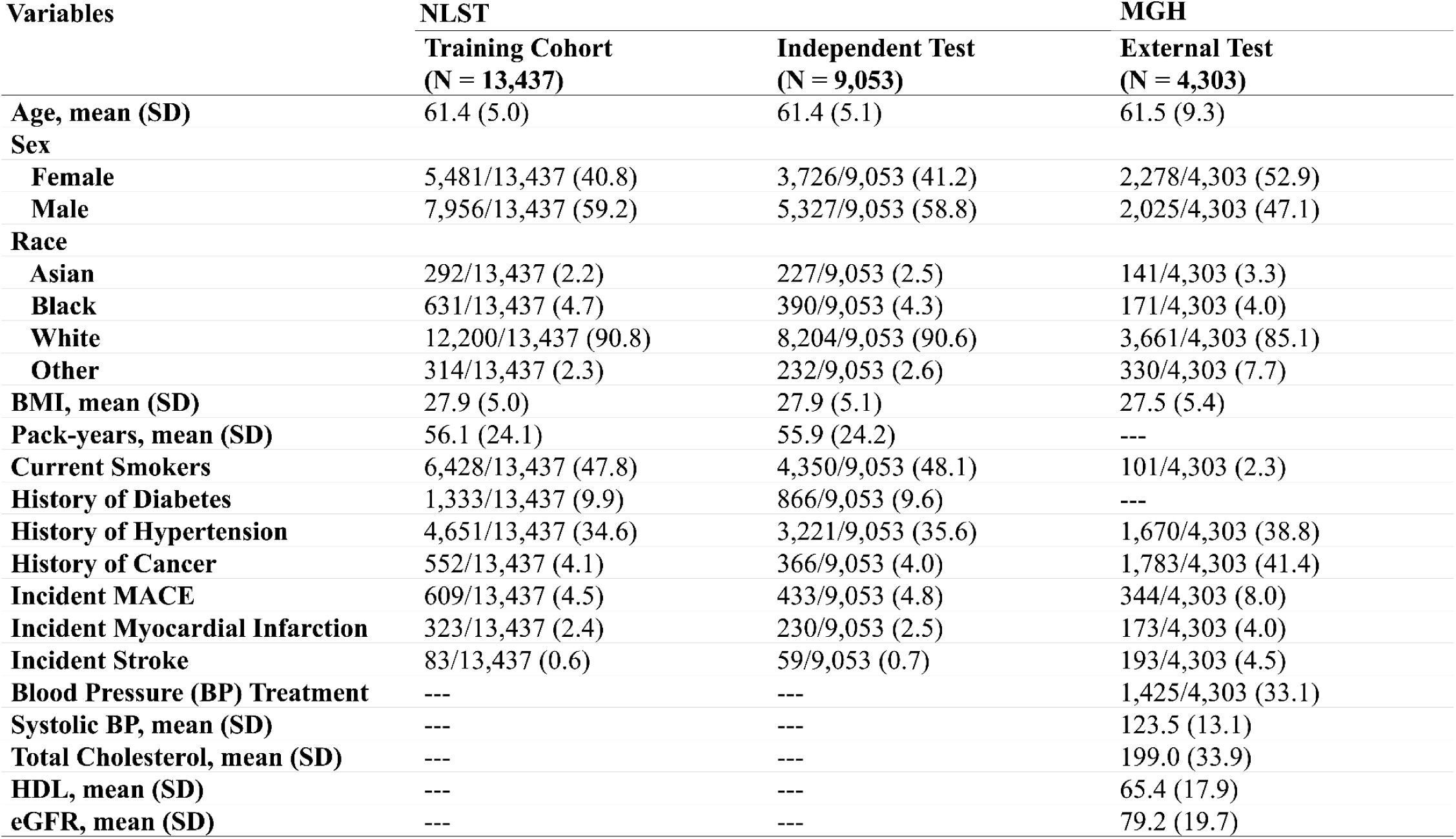
Patient Demographics and Clinical Risk Factors of Training and Testing Cohorts Variables.

We externally tested our model in a cohort of 4,303 individuals who had a non-contrast, non-ECG gated chest CT at a Massachusetts General Hospital (MGH) site and were eligible for primary prevention according to ACC/AHA guidelines (i.e. age 40-75 years, no prior statin usage, no history of MACE, LDL between 70-190 mg/dL, no diabetes) (Table 1). In this cohort, 85.1% (N=3,661) were non-Hispanic white, 47.1% (N=2,025) were male, and the mean age was 61.5 years (SD, 9.3). 4.0% of individuals had incident myocardial infarction and 4.5% of individuals had incident stroke over a median follow-up of 5.1 years. Furthermore, 39.5% (1701/4303) had all inputs available to calculate the PCE risk score, while 30.9% (1328/4303) had all inputs available to calculate the PREVENT risk score.

### Association of Radiomics Features with Cardiovascular Events

We extracted fourteen radiomics features from each of the four cardiac chambers and the aorta, resulting in 70 total features, and conducted an association analysis to assess the association of each radiomics feature with cardiovascular events using Cox proportional hazards regression adjusted for age, sex, and BMI (Supplementary Figures 1-3). We found left ventricular size and shape to be the most predictive of MACE. Per 1 SD, left ventricular volume was associated with a 61% higher risk of fatal MACE, short axis length with 63% higher risk, and sphericity with 32% higher risk. We also found strong associations with aortic surface-to-volume ratio (27% lower risk) and left atrial flatness (45% higher risk).

The radiomics features were then used to train a LASSO regression model to predict 12-year fatal MACE in the NLST training set. The features in the final model and their coefficients are listed in Supplementary Table 3. In stability analysis, we found that these features were robust to bootstrapped resampling. Additionally, test-retest analysis was conducted to measure the reproducibility of each radiomics feature extraction (Supplementary Table 4).

### Discrimination of Risk Scores

We assessed whether the radiomics score could discriminate between those who had or did not have fatal MACE in the independent NLST testing set. Due to the absence of PCE and PREVENT risk score inputs in this cohort, we compared the radiomics model to a baseline regression model trained to predict 12-year MACE using age, sex, race, BMI, pack-years, current smoking, history of diabetes, history of hypertension, and history of cancer (Supplementary Table 10). Addition of radiomics to the baseline regression significantly improved discrimination of fatal MACE (c-index 0.727 vs. 0.707, p < 0.001) as well as discrimination of fatal myocardial infarction (c-index 0.736 vs. 0.700, p < 0.001) but not fatal stroke (Table 2). Calibration analysis of the radiomics score revealed that it was strongly calibrated in males but underestimated risk in females (Supplementary Figure 4).

**Table 2:** Concordance Indices for Risk-Estimation Models (NLST)

| Table 2: Concordance Indices for Risk-Estimation Models (NLST) |  |  |  |
| --- | --- | --- | --- |
| Model | Fatal MACE | Fatal Myocardial Infarction | Fatal Stroke |
| Baseline Regression | 0.707 [0.683, 0.731] | 0.700 [0.667, 0.734] | **0.707** [0.640, 0.774] |
| Radiomics Score | 0.676 [0.649, 0.703] | 0.702 [0.668, 0.736] | 0.511 [0.429, 0.593] |
| Baseline + Radiomics | **0.727** [0.702, 0.752] *** | **0.736** [0.703, 0.768] *** | 0.682 [0.612, 0.752] |

We then tested the discrimination of the radiomics score for 5-year fatal or nonfatal MACE in an external testing dataset of patients who had a routine CT as part of clinical care at a Massachusetts General Hospital (MGH) site. We compared the radiomics score to the PCE and PREVENT risk scores among patients aged 40-75, without diabetes, without prior or current statin usage, and with LDL-C levels between 70-190 mg/dL (patients potentially eligible for primary prevention according to ACC/AHA guidelines). In those who had all inputs available to calculate the PCE risk score (N=1,701; Table 3), the radiomics risk score had greater discrimination than the PCE risk score (c-index 0.640 [0.580, 0.700] vs. 0.567 [0.507, 0.627]), and addition of the radiomics score to the PCE risk score further improved discrimination (c-index 0.653 vs. 0.567, p < 0.001). In the cohort of patients without inputs available to calculate PCE risk (N=2,602), the radiomics risk score had a c-index of 0.592 [0.549, 0.635]. Similar results were found when comparing the radiomics score to the PREVENT score (Table 3).

**Table 3:** Concordance Indices for 5-Year MACE Prediction of Risk-Estimation Models (MGH): All Risk Scores.

|  |  |  |  |  |  |
| --- | --- | --- | --- | --- | --- |
| Model | MGH (All Inputs Available)<br>N = 1328 | MGH (PCE Available)<br>N = 1701 | MGH (PCE Missing)<br>N = 2602 | MGH (PREVENT Available)<br>N = 1328 | MGH (PREVENT Missing)<br>N = 2975 |
| PCE Risk Score | 0.570 [0.495, 0.646] | 0.567 [0.507, 0.627] | NA | NA | NA |
| PREVENT Risk Score | 0.587 [0.513, 0.661] | NA | NA | 0.587 [0.513, 0.661] | NA |
| Radiomics Score | 0.657 [0.590, 0.724] | 0.640 [0.580, 0.700] | **0.592** [0.549, 0.635] | 0.657 [0.590, 0.724] | **0.596** [0.555, 0.636] |
| PCE + Radiomics | 0.671 [0.604, 0.738] *** | **0.653** [0.594, 0.712] *** | NA | NA | NA |
| PREVENT + Radiomics | **0.685** [0.619, 0.751] *** | NA | NA | **0.685** [0.619, 0.751] *** | NA |

**Table 4:** Observed Incident MACE Rates per 1000 Person-Years by Ordinal Radiomics Risk Groups (Rows) and PCE Risk Groups (Columns)

| Radiomics Risk Score | PCE Risk Score |  |  |  |  |
| --- | --- | --- | --- | --- | --- |
|  | Low | Borderline | Intermediate | High | Unknown |
| Low | 6.6 (3.1, 9.8) | 6.1 (1.5, 12.3) | 7.9 (3.2, 13.0) | 23.2 (0.0, 54.6) | 15.8 (12.7, 18.8) |
| Borderline + Intermediate | 15.2 (9.4, 23.4) | 12.4 (6.8, 24.5) | 15.9 (8.3, 15.8) | 15.4 (3.4, 19.1) | 19.4 (15.0, 22.0) |
| High | 102.0 (39.7, 196.4) | 82.3 (32.9, 118.5) | 48.1 (20.3, 76.4) | 50.7 (0.0, 117.9) | 40.0 (24.5, 58.0) |

### Decision Curve Analysis

Decision curve analysis was conducted to quantify the net benefit at various risk thresholds for the PCE risk score, the radiomics risk score, and the combined PCE + Radiomics risk score in comparison to baseline strategies of “treating all” and “treating no” patients (Figure 4). In the cohort of patients with known PCE risk, radiomics had higher net benefit than the PCE risk score across risk thresholds greater than 7.5% (intermediate PCE risk) (Figure 4). The combined risk score had greater net benefit than radiomics alone. In the cohort of patients with unknown PCE risk, the radiomics score had greater net benefit than the “Screen All” regimen past risk thresholds greater than 8%. Similar trends were observed when decision curve analysis was conducted for PREVENT (Supplementary Figure 6).

**Figure 4:**
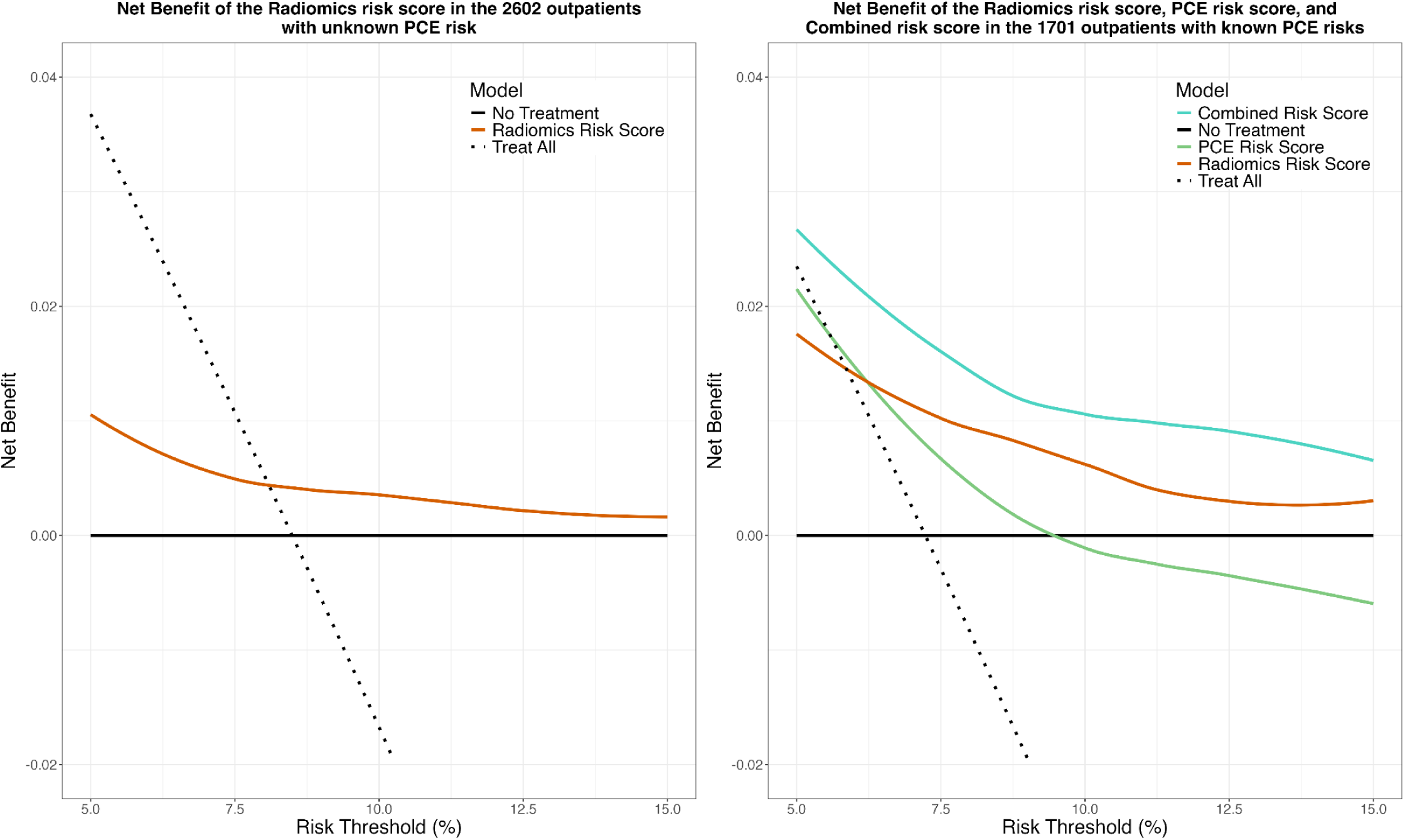
Decision curve analysis for the Radiomics risk score, PCE risk score, and Combined risk score.

### Risk Stratification

We then stratified the radiomics score into four ordinal groups (low (<3.0%), borderline (3.0% to <5.0%), intermediate (5.0% to <7.5%), high (≥7.5%)) and assessed the association of radiomics and PCE ordinal groups with incident MACE. Among those with inputs available to calculate PCE risk (N=1,701), cumulative incidence curves demonstrated a graded association of radiomics risk groups with incident MACE, and these groups better stratified risk than standard PCE risk groups (Figure 3). The radiomics groups also stratified risk in those with inputs missing to calculate PCE risk (N=2,602).

We next assessed risk reclassification between the radiomics risk score and the PCE risk score. Among patients at low PCE risk, radiomics categories had a graded association with MACE (6.6 - 102.0 incident MACE per 1000 person-years from low to high radiomics risk). Similar results were found for patients at borderline (6.1 - 82.3 incident MACE per 1000 person-years) and intermediate (7.9 - 48.1 incident MACE per 1000 person-years) PCE risk. However, this association was not seen in the high PCE risk group. Similar results were found when comparing the radiomics categories to PREVENT risk categories (Supplementary Table 8).

### Statin Eligibility

We then compared rates of incident MACE per 1000 person-years among patients considered “statin-eligible” vs. “ineligible” using a single binary threshold delineating low/borderline from intermediate/high risk by the radiomics and PCE risk scores (Table 5). The 11.8% of individuals (200/1701) that were statin-eligible by both scores had a 2.6-fold higher rate of incident MACE than those that were statin-eligible by the PCE risk score alone (29.5 [20.5, 39.1] vs. 11.2 [8.0, 14.4] incident MACE per 1000 person-years). The radiomics risk score reclassified 69 patients as statin-eligible (38.3 incident MACE per 1000 person-years) and 646 patients as ineligible (11.2 incident MACE per 1000 person-years). In patients missing inputs to calculate PCE risk, statin-eligible patients had a 1.8-fold higher incidence of MACE than ineligible patients by the radiomics risk score (29.5 [21.9, 37.6] vs. 16.7 [14.3, 19.0] incident MACE per 1000 person-years). Similar results were seen when comparing to the PREVENT score (Supplementary Table 9).

**Table 5:** Observed Incident MACE Rates per 1000 Person-Years by Statin Eligibility Groups Defined by the Radiomics Risk Score (Rows) and the PCE Risk Score (Columns)

| Statin Eligibility by Radiomics Risk Score | Statin Eligibility by PCE Risk Score |  |  |
| --- | --- | --- | --- |
|  | Risk < 7.5% | Risk ≥ 7.5% | PCE Missing |
| Risk < 5.0% | 9.0 (6.4, 11.9) | 11.2 (8.0, 14.4) | 16.7 (14.3, 19.0) |
| Risk ≥ 5.0% | 38.3 (20.7, 59.2) | 29.5 (20.5, 39.1) | 29.5 (21.9, 37.6) |

## DISCUSSION

Accurate assessment of cardiovascular risk is the foundation of primary prevention. In this study, we tested whether structural phenotypes of the heart and aorta, measured automatically via deep learning from non-contrast chest CT, predicted cardiovascular risk beyond known cardiovascular risk factors. We developed a regression model that predicted an individual’s 12-year risk of fatal MACE using radiomics features extracted with an open-source segmentation tool (8) from a single chest CT. We then compared this “radiomics score” to established cardiovascular risk factors in a lung cancer screening population and to the clinical standard PCE and PREVENT risk scores in an external validation set of 4,303 patients who had routine clinical non-contrast enhanced chest CT and were potentially eligible for primary prevention according to guidelines from the ACC/AHA.

Our main findings are as follows. First, in the internal NLST testing cohort, we found that the radiomics score provided added value to baseline cardiovascular risk factors, significantly improving discrimination of fatal MACE (c-index 0.727 vs. 0.707, p < 0.001). Second, we compared the radiomics score to the clinical standard PCE and PREVENT risk scores in an external testing set from Massachusetts General Hospital. We found that the radiomics score had better discrimination than the PCE risk score (radiomics c-index 0.640 [0.580, 0.700] vs. PCE c-index 0.567 [0.507, 0.627]) as well as the PREVENT risk score (radiomics c-index 0.657 [0.590, 0.724] vs. PREVENT c-index 0.587 [0.513, 0.661]). Third, we stratified the continuous radiomics score into ordinal risk groups and found that these groups had a graded association with incident MACE in both patients with inputs available to calculate PCE risk (6.6 - 102.0 incident MACE per 1000 person-years in low PCE risk group; 6.1 - 82.3 in borderline PCE risk group; 7.9 - 48.1 in intermediate PCE risk group) and in patients with unknown PCE risk (60% of cohort; 15.8 - 40.0 incident MACE per 1000 person-years). Last, we determined that certain phenotypes of heart shape, such as left ventricular volume and sphericity, aortic surface to volume ratio, and left atrial flatness, were strongly associated with risk of future MACE.

Our results suggest that a radiomics approach can improve risk assessment by leveraging currently unused information from routine chest CT, including in patients undergoing lung cancer screening. We envision the radiomics tool being useful in an opportunistic screening paradigm, where the tool is automatically applied to existing CT exams in the electronic medical record. We found that the radiomics score improved risk estimation in those who had all risk factors to calculate existing PCE and PREVENT risk scores; however, it also accurately identified high-risk patients in the 60% of patients with at least one missing input to these calculators. In patients with known PCE or PREVENT risk, the radiomics score could serve as a “risk-enhancing” factor to further stratify risk. In patients missing inputs to the PCE risk score but with an existing chest CT, the model could be used to identify patients that may benefit from risk factor assessment. The model can be efficiently applied to existing CTs in the background of the medical record (<2 min/CT on a standard computer) to calculate a patient’s risk with little disruption to the clinical workflow.

Our radiomics model combines 13 structural phenotypes to assess cardiovascular risk: aortic size (diameter of the largest axial slice, short axis length), aortic shape (surface-volume ratio), left atrial size (short axis length), left atrial shape (flatness, sphericity), left ventricular size (volume, minor axis length), left ventricular sphericity, right atrial shape (major axis length), right atrial sphericity, and right ventricular size (short axis length and surface-volume ratio). These features were identified as the most predictive in our development dataset and were stable across bootstrapped versions of the dataset. Most selected phenotypes have well-established associations with cardiovascular risk, suggesting biological plausibility. Size of the ascending and descending aorta are associated with prevalent cardiovascular risk factors (BMI, hypertension, diabetes) (26) and cardiovascular mortality in a sex-specific manner (27); however, the association of aortic shape with atherosclerotic cardiovascular disease is not well understood. Cardiac chamber size, including the left atrium (28,29), the left ventricle (30,31), and the right ventricle (32) have well-established associations with cardiovascular risk, and left ventricular sphericity has been demonstrated to be a strong predictor (11).

Limitations of our study must be addressed before deployment. Our study is retrospective; prospective clinical trials are necessary to establish the clinical utility of this automated risk scoring system. Second, the model was trained on clinical trial participants undergoing lung cancer screening CT, and it was trained to predict only *fatal* MACE. For external testing, we included non-fatal MACE, as this is used by the PCE and PREVENT risk calculators. Additionally, the external testing cohort consisted of patients at intermediate LDL-C and not on a statin, the population on which the PCE and PREVENT calculators would be applied. This criteria could not be applied to the NLST cohort due to unavailable data, resulting in miscalibration that could be corrected with further fine-tuning including non-fatal outcomes. Third, the LASSO model identified features associated with increased risk, but does not provide a physiologic explanation linking these features with MACE. This opens future investigation for studying the role of heart and aortic structure in cardiac physiology and cardiovascular disease etiology. A benefit of the radiomics approach is that the features are inherently interpretable; however, less interpretable deep learning-derived features may further improve risk estimation (33) and will be investigated in future work. Fourth, our training and testing cohorts were largely non-Hispanic white, and our model needs to be validated in more diverse cohorts to ensure generalizability. Finally, future studies will need to evaluate radiomics in the context of the coronary artery calcium score, which can also be derived from chest CT and has been shown to be predictive of MACE (10,12).

In conclusion, we developed an interpretable machine learning model that predicts cardiovascular risk using quantitative, structural features of the heart and aorta. This “radiomics” approach improves on existing clinical risk estimation algorithms and sheds light on cardiac and aortic phenotypes linked to MACE. This may enable opportunistic cardiovascular risk assessment from routine chest CT to guide primary prevention of cardiovascular disease.

## Data Availability

All data produced in the present study are available upon reasonable request to the authors.

