## Supplemental Figures and Tables for "OPPORTUNISTIC ASSESSMENT OF CARDIOVASCULAR RISK USING AI-DERIVED STRUCTURAL AORTIC AND CARDIAC PHENOTYPES FROM NON-CONTRAST CHEST COMPUTED TOMOGRAPHY"

### SUPPLEMENT CONTENTS


#### I Supplementary Results

- Risk stratification of the radiomics risk score and PREVENT risk score
- Statin eligibility by the radiomics risk score and PREVENT risk score
- Decision curve analysis for the radiomics risk score and PREVENT risk score in cohorts of patients with known and unknown PREVENT risk

#### II Supplementary Figures

- Supplementary Figure 1: Association of Individual Radiomics Features with Fatal MACE
- Supplementary Figure 2: Association of Individual Radiomics Features with Fatal Myocardial Infarction
- Supplementary Figure 3: Association of Individual Radiomics Features with Fatal Stroke
- Supplementary Figure 4: Calibration plots for Radiomics Risk Score to Estimate 12-year MACE Risk Stratified by Gender
- Supplementary Figure 5: Cumulative Incidence Curves for the Radiomics Risk Score and the PREVENT Risk Score
- Supplementary Figure 6: Decision Curve Analysis for the Radiomics Risk Score, PREVENT Risk Score, and Combined Risk Score

#### III Supplementary Tables

- Supplementary Table 1: International Classification of Diseases Codes for Cardiovascular Mortality and MACE
- Supplementary Table 2: List and Definitions of Radiomics Features
- Supplementary Table 3: LASSO Radiomics Model Variables and Coefficients
- Supplementary Table 4: Test-Retest Analysis of Radiomics Features
- Supplementary Table 5: Stability Analysis of Radiomics Features for Prediction of Fatal MACE
- Supplementary Table 6: Stability Analysis of Radiomics Features for Prediction of Fatal Myocardial Infarction
- Supplementary Table 7: Stability Analysis of Radiomics Features for Prediction of Fatal Stroke
- Supplementary Table 8: Observed Incident MACE per 1000 Person-Years and Risk Classification Using Radiomics Risk Score versus PREVENT Score

- Supplementary Table 9: Observed Incident MACE per 1000 Person-Years and Statin Eligibility Using Radiomics Risk Score versus PREVENT Score
- Supplementary Table 10: NLST Baseline Model Variables

### I. SUPPLEMENTARY RESULTS

**Decision Curve Analysis for the Radiomics Risk Score and PREVENT Risk Score in Cohorts of Patients with Unknown and Known PREVENT Risk**

Decision curve analysis was conducted to compare the net benefit of the radiomics risk score, the PREVENT risk score, and a combined PREVENT + Radiomics risk score. In the cohort of patients with known PREVENT risk, radiomics had higher net benefit than the PREVENT risk score across risk thresholds greater than 8% (Supplementary Figure 6). The combined risk score had greater net benefit than both radiomics and PREVENT alone. In the cohort of patients with unknown PREVENT risk, the radiomics score had greater net benefit than the treat all regimen past risk thresholds greater than 7.5%.

**Distribution of Radiomics Risk Score vs. PREVENT Risk Score and Reclassification of Incident MACE:**

We assessed the association of radiomics and PREVENT ordinal groups with incident MACE. Among those with inputs available to calculate PREVENT risk (N=1,328), cumulative incidence curves demonstrated a graded association of radiomics with incident MACE and showed better stratification with the radiomics score than with the standard PREVENT risk groups (Supplementary Figure 5). The radiomics score also stratified risk in those with inputs missing to calculate PREVENT risk (N=2,975).

We next analyzed risk reclassification between the radiomics risk score and the PREVENT risk score. There were no patients classified as high risk by the PREVENT risk score. For low, borderline, and intermediate PREVENT risk categories, the radiomics score displayed a graded association with MACE (4.5 - 92.8 incident MACE per 1000 person-years from low to high radiomics risk in the low PREVENT group; 7.8 - 48.6 in the borderline PREVENT group; 13.3 - 39.4 in the intermediate PREVENT group) (Supplementary Table 8).

**Observed Incident MACE and Statin Eligibility Using Radiomics Risk Score versus PREVENT Score**

We compared rates of incident MACE per 1000 person-years using a single binary threshold delineating low/borderline from intermediate/high risk by the radiomics and PREVENT risk scores. The 6.6% of individuals (87/1328) that were at high-risk by both scores had a 2.1-fold higher incidence of MACE than those that were at high-risk by the PREVENT score alone (32.1 [19.1, 45.7] vs. 15.1 [8.7, 21.8] incident MACE per 1000 person-years). The radiomics risk score reclassified 134 patients as high-risk (28.4 incident MACE per 1000 person-years) and 207 patients as low-risk (15.1 incident MACE per 1000 person-years). In patients missing inputs to calculate PREVENT risk, statin-eligible patients had a 1.9-fold higher incidence of MACE than statin-ineligible patients by the radiomics risk score (30.8 [23.5, 38.5] vs. 16.3 [14.1, 18.6] incident MACE per 1000 person-years).

### II. SUPPLEMENTARY FIGURES

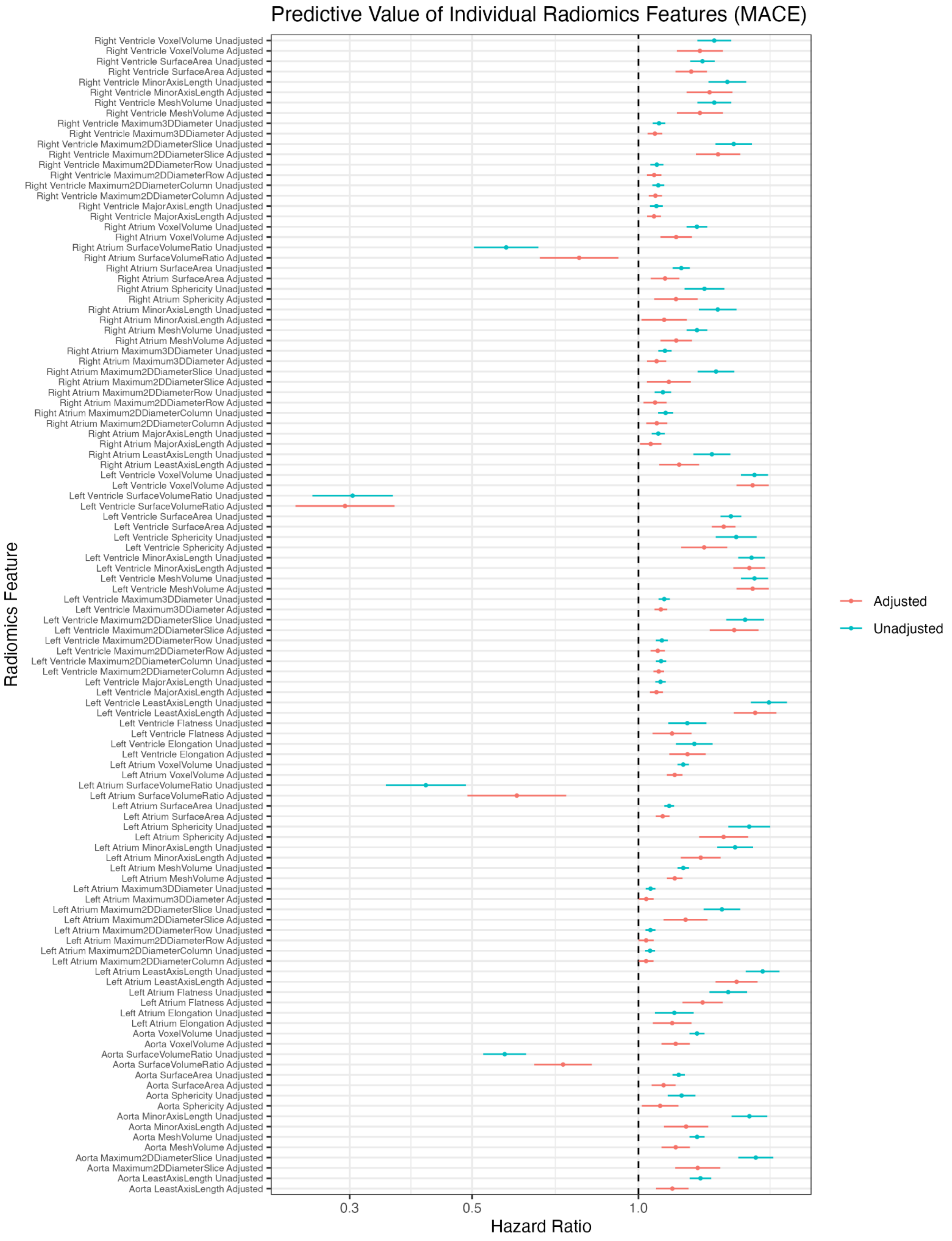


**Supplementary Figure 1: Association of Individual Radiomics Features with Fatal MACE**

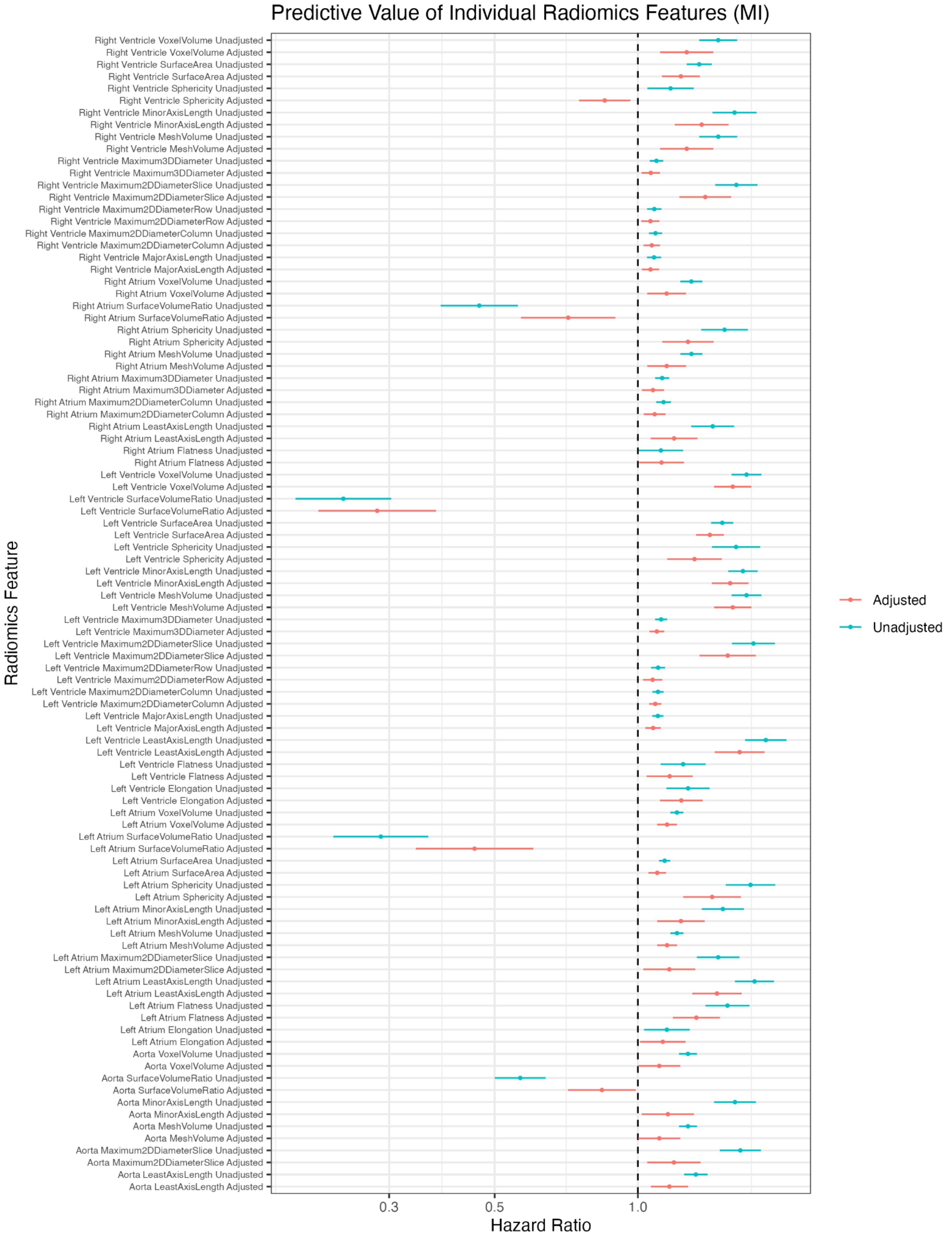


**Supplementary Figure 2: Association of Individual Radiomics Features with Fatal Myocardial Infarction**

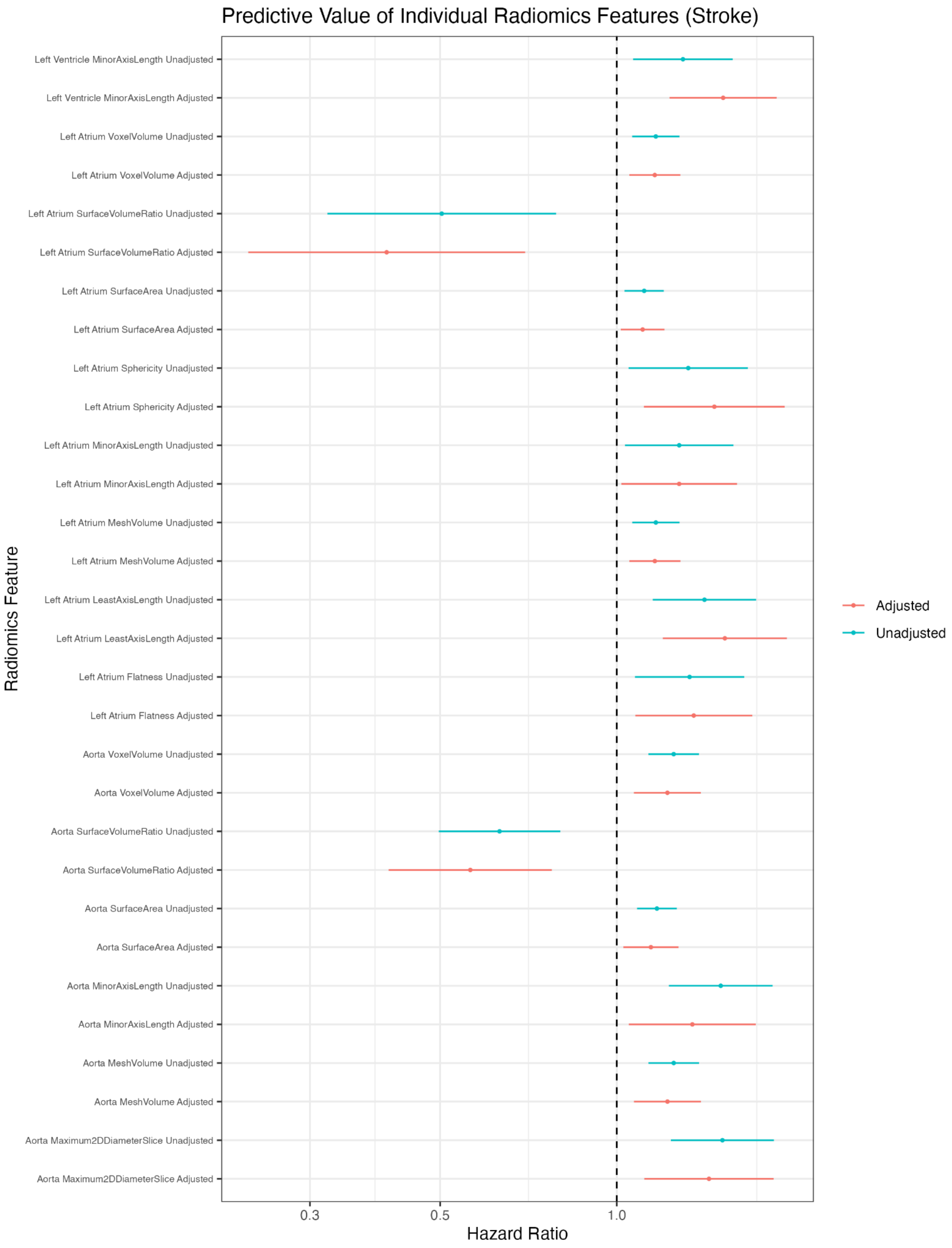


**Supplementary Figure 3: Association of Individual Radiomics Features with Fatal Stroke**

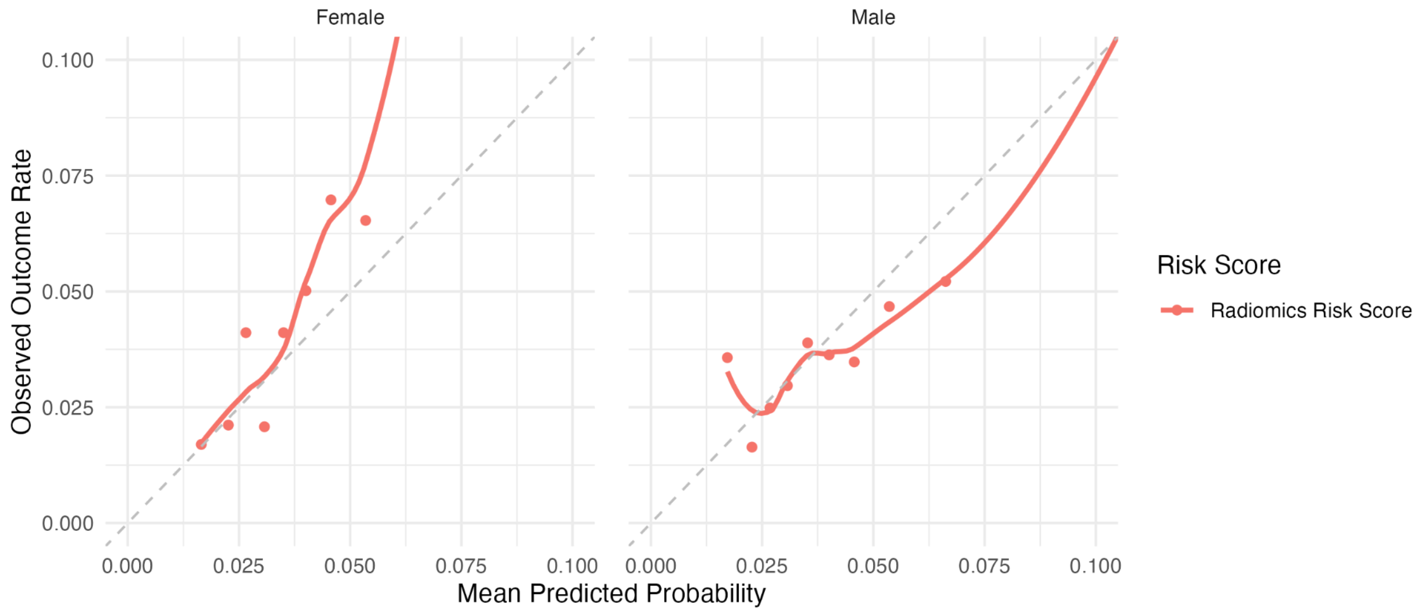


**Supplementary Figure 4: Calibration plots for Radiomics Risk Score to Estimate 12-year MACE Risk Stratified by Gender**

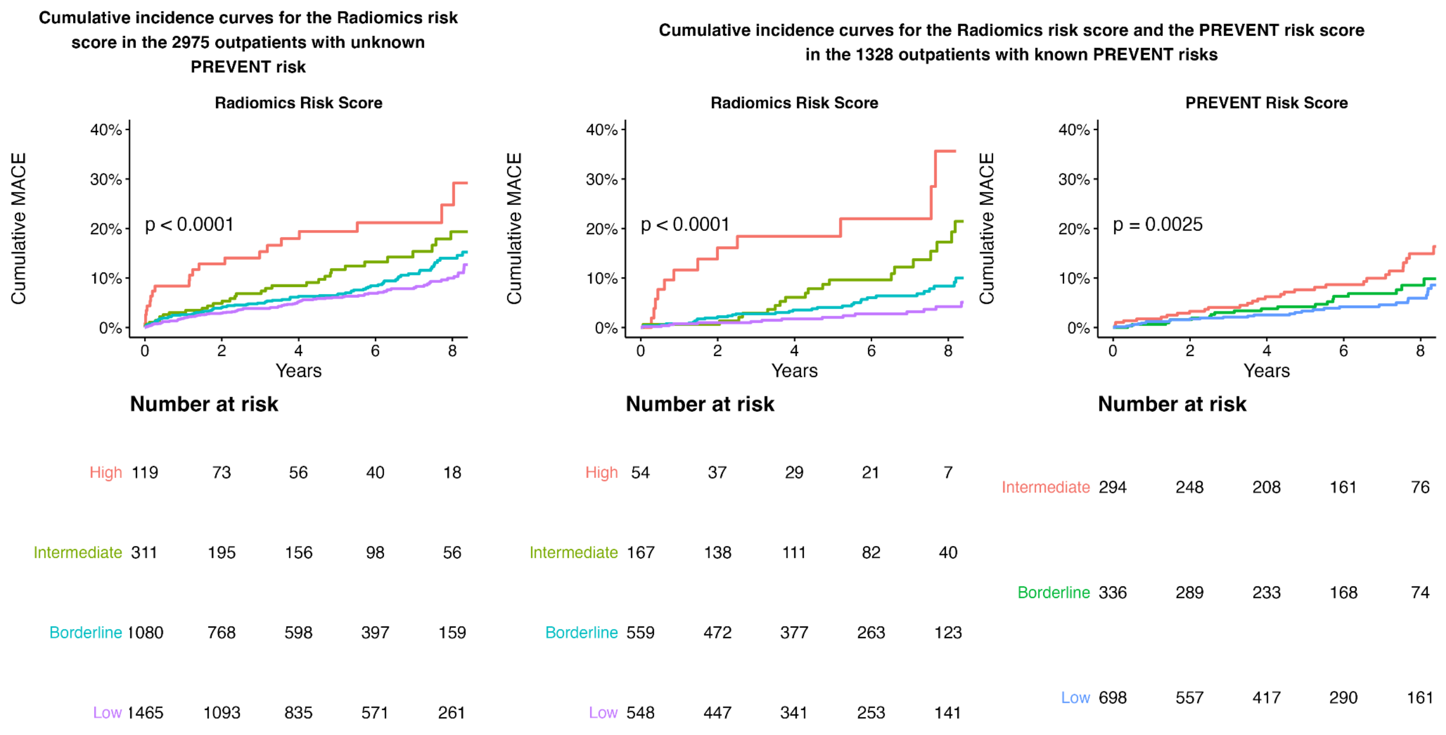


**Supplementary Figure 5: Cumulative Incidence Curves for the Radiomics Risk Score and the PREVENT Risk Score**

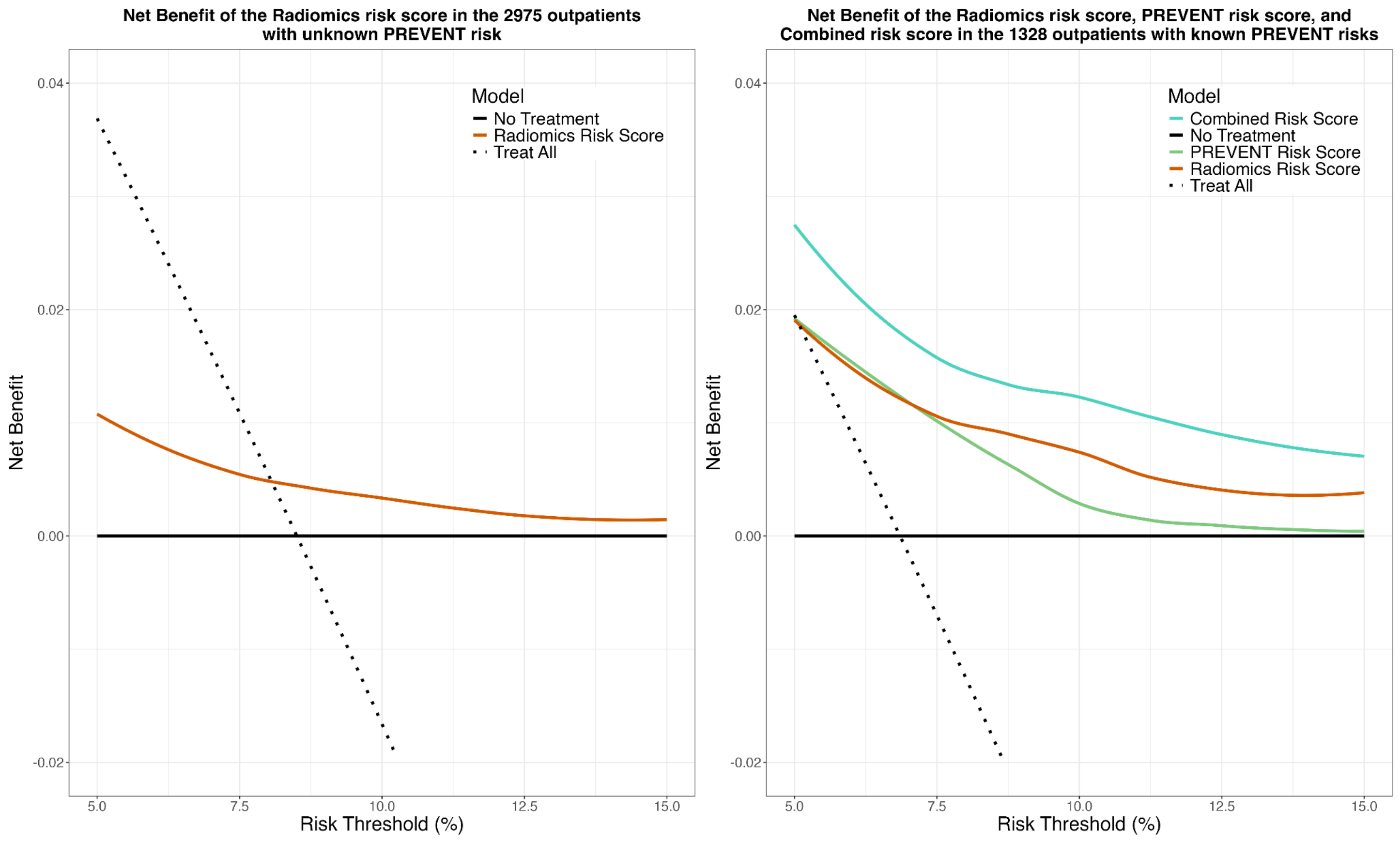


**Supplementary Figure 6: Decision Curve Analysis for the Radiomics Risk score, PREVENT Risk score, and Combined Risk Score.**

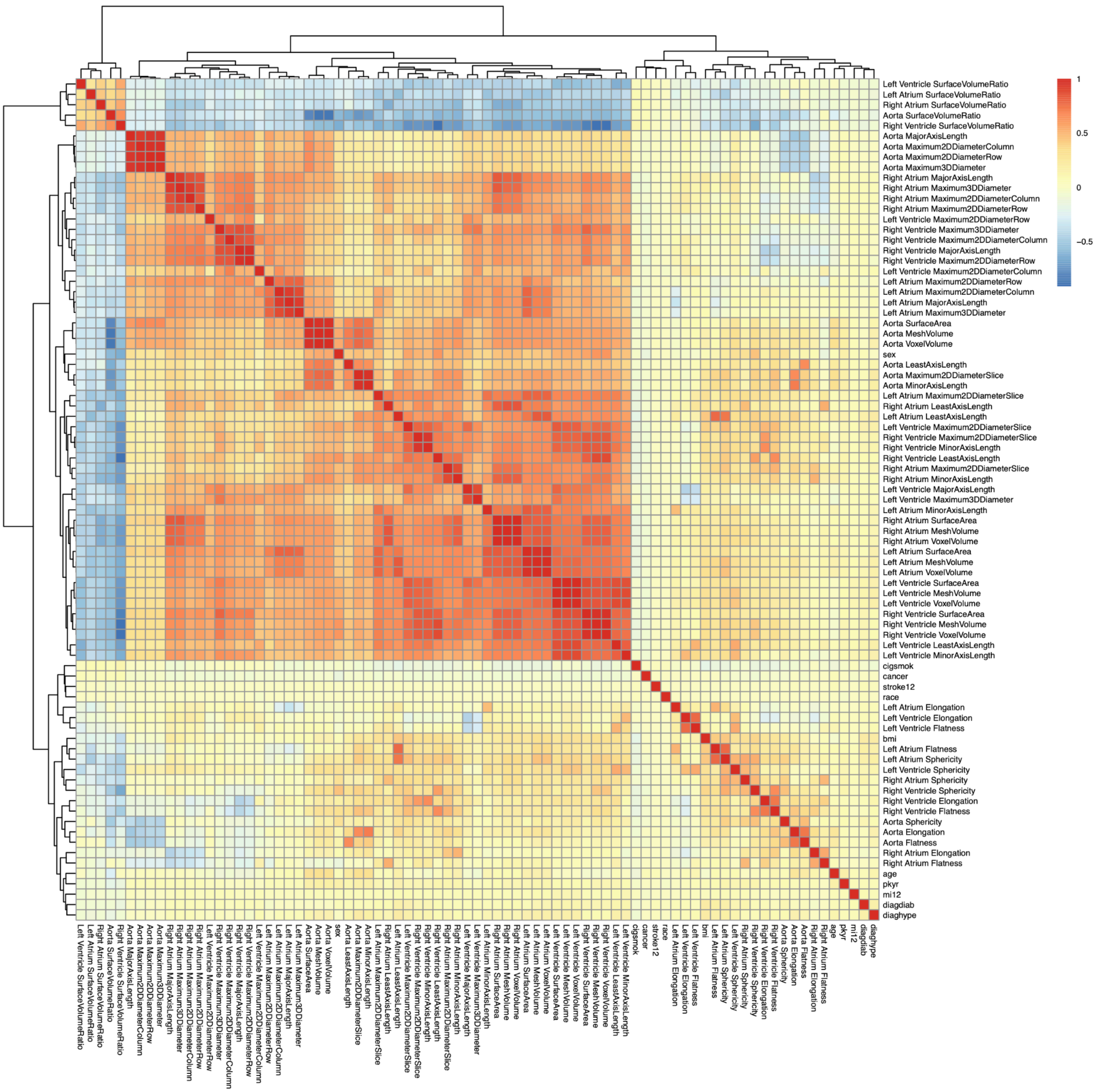


**Supplementary Figure 7: Concordance Matrix of Radiomics Features**

### III. SUPPLEMENTARY TABLES

**Supplementary Table 1:** International Classification of Diseases Codes for cardiovascular disease-related mortality (training cohort) and incident major adverse cardiovascular events (testing cohort)

| | **ICD9** | **ICD10** |
|---|---|---|
| | ***Training cohort*** | |
| | None | I1*, I2*, I4*, I50, I51, I52, I6*, I7* |
| | ***Testing cohort*** | |
| **Stroke** | 433.11, 433.01, 433.21, 433.31, 433.81, 433.91, 434.01, 434.11, 434.91 | I63* |
| **Myocardial Infarction** | 36.1, 36.2, 411 | I21, I22, I23 |
| **History of Type 2 Diabetes** | 249, 249.01, 249.1, 249.11, 249.2, 249.21, 249.21, 249.3, 249.31, 249.4, 249.41, 249.5, 249.51, 249.6, 249.61, 249.7, 249.71, 249.8, 249.81, 249.9, 249.91, 250, 250.01, 250.02, 250.03, 250.1, 250.11, 250.12 250.13, 250.2, 250.21, 250.22, 250.23, 250.3, 250.31, 250.32, 250.33, 250.4, 250.41, 250.42, 250.43, 250.5, 250.51, 250.52, 250.53, 250.6, 250.61, 250.62, 250.63, 250.7, 250.71, 250.72, 250.73, 250.8, 250.81, 250.82, 250.83, 250.9, 250.91, 250.92, 250.93, 357.2, 362.01, 362.02, 362.03, 362.04, 362.05, 362.06, 362.07, 366.41 | E08.00,E08.01,E08.10, E08.11,E08.21,E08.22, E08.29,E08.31,E08.32, E08.33,E08.34,E08.35, E08.36,E08.39,E08.40, E08.41,E08.42,E08.43, E08.44,E08.49,E08.51, E08.52,E08.61,E08.62, E08.63,E08.64,E08.65, E08.69,E08.8,E08.9,E09.00, E09.01,E09.10,E09.11, E09.21,E09.22,E09.29, E09.31,E09.32,E09.33, E09.34,E09.35,E09.36, E09.39,E09.40,E09.41, E09.42,E09.43,E09.44, E09.49,E09.51,E09.52, E09.59,E09.61,E09.62, E09.63,E09.64,E09.65, E09.69,E09.8,E09.9,E09.10, E09.11,E09.21,E09.22, E09.29,E09.31,E09.32, E09.33,E09.34,E09.35, E09.36,E09.39,E09.40, E09.41,E09.42,E09.43, E09.44,E09.49,E09.51, |

| | | |
|---|---|---|
| | | E09.52,E09.59,E09.61,E09.62,E09.63,E09.64,E09.65, E09.69,E09.8,E09.9,E11.00, E11.01,E11.21,E11.22,E11.29,E11.31,E11.32,E11.33, E11.34,E11.35,E11.36, E11.39,E11.40,E11.41,E11.42,E11.51,E11.52,E11.59, E11.61,E11.62,E11.63,E11.64,E11.65,E11.69,E11.8,E11.9, E13.00,E13.11,E13.21, E13.22,E13.29,E13.31, E13.32,E13.33,E13.34, E13.35,E13.36,E13.39, E13.40,E13.41,E13.42, E13.43,E13.44,E13.49, E13.51,E13.52,E13.59, E13.61,E13.62,E13.63, E13.64,E13.65,E13.69, E13.8,E13.9,R82.4 |

International Classification of Diseases Codes for cardiovascular disease-related mortality and incident major adverse cardiovascular events.
ICD=International Classification of Diseases Codes

**Supplementary Table 2: Radiomics Features**

| Radiomics Feature | Formula | Description |
|---|---|---|
| Elongation | Elongation = $\sqrt{\frac{\lambda_{minor}}{\lambda_{major}}}$ | Elongation is computed from the lengths of the largest and second largest principle components of the region of interest. The value ranges from 0 to 1, in which an elongation of 1 describes a cross section that is circular. An elongation of 0 describes a cross section that is |

| | | |
|---|---|---|
| | | linear. |
| Flatness | Flatness = $\sqrt{\frac{\lambda_{least}}{\lambda_{major}}}$ | Flatness is computed from the lengths of the largest and smallest principle components of the region of interest. The value ranges from 0 to 1, in which a flatness of 1 describes a sphere-like object and a flatness of 0 describes a flat object. |
| Least Axis Length | Least Axis = $4\sqrt{\lambda_{least}}$ | Least Axis Length describes the smallest axis of the ROI-enclosing ellipsoid. It is computed from the largest principle component. |
| Minor Axis Length | Minor Axis = $4\sqrt{\lambda_{minor}}$ | Minor Axis Length describes the second largest axis of the ROI-enclosing ellipsoid. It is computed from the second largest principle component. |
| Major Axis Length | Major Axis = $4\sqrt{\lambda_{major}}$ | Major Axis Length describes the largest axis of the ROI-enclosing ellipsoid. It is computed from the largest principle component. |
| Maximum 2D Diameter Slice | Euclidean Distance | Maximum 2D Diameter Slice is computed as the largest Euclidean |

| | | |
|---|---|---|
| | | distance between surface mesh vertices in the axial plane. |
| Maximum 2D Diameter Column | Euclidean Distance | Maximum 2D Diameter Column is computed as the largest Euclidean distance between surface mesh vertices in the coronal plane. |
| Maximum 3D Diameter | Euclidean Distance | Maximum 3D Diameter is computed as the largest Euclidean distance between surface mesh vertices. |
| Surface Area | Surface Area = $\sum_{i=1}^{N} \frac{1}{2} \left\| a_i b_i \times a_i c_i \right\|$ | Surface Area is computed by summing the areas of the triangles formed by the vertices of the surface mesh. |
| Mesh Volume | Mesh Volume = $\sum_{i=1}^{N} \frac{Oa_i \cdot (Ob_i \times Oc_i)}{6}$ | Surface Area is computed by summing the volumes of the tetrahedrons formed by the vertices of the triangle mesh. |
| Sphericity | Sphericity = $\frac{\sqrt[3]{36\pi V^2}}{A}$ | Sphericity computes the roundness of the segmented ROI relative to a sphere. Sphericity ranges from 0 to 1, with 1 indicating a perfect sphere. |

| | | |
|---|---|---|
| Surface Volume Ratio | Surface to Volume Ratio = $\frac{A}{V}$ | Surface to Volume Ratio computes how compact an object is. A small value indicates a highly compact object. |

**Supplementary Table 3: Radiomics Model Variables**

| Radiomics Feature | Stability | Model Coefficient |
|---|---|---|
| Aorta LeastAxisLength | 100% | 0.037 |
| Aorta Maximum2DDiameterSlice | 100% | 0.982 |
| Aorta SurfaceVolumeRatio | 100% | -0.202 |
| Left Atrium Flatness | 85.6% | 0.044 |
| Left Atrium LeastAxisLength | 100% | 0.126 |
| Left Atrium Sphericity | 100% | 0.055 |
| Left Ventricle MeshVolume | 100% | 0.266 |
| Left Ventricle MinorAxisLength | 100% | 0.141 |
| Left Ventricle Sphericity | 100% | 0.040 |
| Right Atrium MajorAxisLength | 100% | -0.015 |
| Right Atrium Sphericity | 99.9% | 0.005 |
| Right Ventricle LeastAxisLength | 99.5% | -0.166 |
| Right Ventricle SurfaceVolumeRatio | 99.2% | 0.149 |

#### Supplementary Table 4: Test-Retest Analysis

| Radiomics Feature | Concordance Correlation Coefficient (CCC) | Intraclass Correlation Coefficient (ICC) |
|---|---|---|
| Aorta Elongation | 0.831102811 | 0.831120235 |
| Aorta Flatness | 0.820833235 | 0.820851491 |
| Aorta LeastAxisLength | 0.973386402 | 0.973389618 |
| Aorta MajorAxisLength | 0.649126708 | 0.649154981 |
| Aorta Maximum2DDiameterColumn | 0.720806113 | 0.720831095 |
| Aorta Maximum2DDiameterRow | 0.661319149 | 0.661346952 |
| Aorta Maximum2DDiameterSlice | 0.964280001 | 0.964284277 |
| Aorta Maximum3DDiameter | 0.669992564 | 0.670020011 |
| Aorta MeshVolume | 0.932223742 | 0.932231585 |
| Aorta MinorAxisLength | 0.960551665 | 0.960556368 |
| Aorta Sphericity | 0.680464426 | 0.680491418 |
| Aorta SurfaceArea | 0.889370439 | 0.889382652 |
| Aorta SurfaceVolumeRatio | 0.923113761 | 0.923122571 |
| Aorta VoxelVolume | 0.932502984 | 0.932510797 |
| Left Atrium Elongation | 0.921008217 | 0.921017247 |
| Left Atrium Flatness | 0.944922588 | 0.944929048 |
| Left Atrium LeastAxisLength | 0.962837551 | 0.962841992 |
| Left Atrium MajorAxisLength | 0.6460356 | 0.646063987 |
| Left Atrium Maximum2DDiameterColumn | 0.611093175 | 0.611122678 |
| Left Atrium Maximum2DDiameterRow | 0.601185875 | 0.601215639 |
| Left Atrium Maximum2DDiameterSlice | 0.925977184 | 0.925985693 |
| Left Atrium Maximum3DDiameter | 0.500308134 | 0.500339169 |
| Left Atrium MeshVolume | 0.935656126 | 0.935663599 |
| Left Atrium MinorAxisLength | 0.924759107 | 0.924767744 |
| Left Atrium Sphericity | 0.803872167 | 0.803891739 |
| Left Atrium SurfaceArea | 0.877004877 | 0.877018267 |
| Left Atrium SurfaceVolumeRatio | 0.15818616 | 0.158202691 |
| Left Atrium VoxelVolume | 0.935619094 | 0.935626571 |
| Left Ventricle Elongation | 0.958437419 | 0.958442364 |
| Left Ventricle Flatness | 0.924135998 | 0.9241447 |
| Left Ventricle LeastAxisLength | 0.906101265 | 0.906111826 |
| Left Ventricle MajorAxisLength | 0.832736332 | 0.832753623 |
| Left Ventricle Maximum2DDiameterColumn | 0.803941318 | 0.803960884 |
| Left Ventricle Maximum2DDiameterRow | 0.845305695 | 0.845321927 |
| Left Ventricle Maximum2DDiameterSlice | 0.973232402 | 0.973235636 |
| Left Ventricle Maximum3DDiameter | 0.835484355 | 0.835501417 |
| Left Ventricle MeshVolume | 0.963297513 | 0.963301902 |
| Left Ventricle MinorAxisLength | 0.937279916 | 0.937287213 |
| Left Ventricle Sphericity | 0.6848829 | 0.684909691 |
| Left Ventricle SurfaceArea | 0.940822874 | 0.940829785 |
| Left Ventricle SurfaceVolumeRatio | 0.312539031 | 0.312565704 |
| Left Ventricle VoxelVolume | 0.963395219 | 0.963399596 |
| Right Atrium Elongation | 0.953292295 | 0.953297822 |
| Right Atrium Flatness | 0.933214914 | 0.933222651 |
| Right Atrium LeastAxisLength | 0.972245634 | 0.972248984 |
| Right Atrium MajorAxisLength | 0.810190539 | 0.810209629 |
| Right Atrium Maximum2DDiameterColumn | 0.828937039 | 0.828954642 |
| Right Atrium Maximum2DDiameterRow | 0.852939398 | 0.852954969 |
| Right Atrium Maximum2DDiameterSlice | 0.979627394 | 0.979629871 |
| Right Atrium Maximum3DDiameter | 0.840092074 | 0.84010875 |
| Right Atrium MeshVolume | 0.956078433 | 0.956083646 |
| Right Atrium MinorAxisLength | 0.95439806 | 0.954403463 |
| Right Atrium Sphericity | 0.808467818 | 0.80848704 |
| Right Atrium SurfaceArea | 0.926829106 | 0.926837524 |
| Right Atrium SurfaceVolumeRatio | 0.649884294 | 0.64991254 |
| Right Atrium VoxelVolume | 0.95610738 | 0.956112589 |
| Right Ventricle Elongation | 0.960386051 | 0.960390773 |
| Right Ventricle Flatness | 0.954981506 | 0.954986842 |
| Right Ventricle LeastAxisLength | 0.97199684 | 0.972000219 |
| Right Ventricle MajorAxisLength | 0.766204651 | 0.766226888 |
| Right Ventricle Maximum2DDiameterColumn | 0.770185163 | 0.770207135 |
| Right Ventricle Maximum2DDiameterRow | 0.746008161 | 0.746031682 |
| Right Ventricle Maximum2DDiameterSlice | 0.968225011 | 0.96822883 |
| Right Ventricle Maximum3DDiameter | 0.753281684 | 0.753304754 |
| Right Ventricle MeshVolume | 0.969657839 | 0.969661491 |
| Right Ventricle MinorAxisLength | 0.960601452 | 0.96060615 |
| Right Ventricle Sphericity | 0.870527318 | 0.870541309 |
| Right Ventricle SurfaceArea | 0.934221187 | 0.934228815 |
| Right Ventricle SurfaceVolumeRatio | 0.946369847 | 0.946376147 |
| Right Ventricle VoxelVolume | 0.969645142 | 0.969648795 |

##### Supplementary Table 5: Most Stable Radiomic Features for Predicting MACE

| Radiomics Feature | Stability | Adjusted Hazard Ratio (CI) |
|---|---|---|
| Aorta LeastAxisLength | 100% | 1.15 (1.08, 1.23) |
| Aorta Maximum2DDiameterSlice | 100% | 1.28 (1.17, 1.41) |
| Aorta SurfaceVolumeRatio | 100% | 0.73 (0.65, 0.82) |
| Left Atrium LeastAxisLength | 100% | 1.51 (1.38, 1.64) |
| Left Atrium Sphericity | 100% | 1.43 (1.29, 1.58) |
| Left Ventricle MeshVolume | 100% | 1.61 (1.51, 1.72) |
| Left Ventricle MinorAxisLength | 100% | 1.59 (1.49, 1.70) |
| Left Ventricle Sphericity | 100% | 1.32 (1.19, 1.45) |
| Right Atrium MajorAxisLength | 100% | 1.05 (1.01, 1.10) |
| Right Atrium Sphericity | 99.9% | 1.17 (1.07, 1.28) |
| Right Ventricle LeastAxisLength | 99.5% | 1.02 (0.92, 1.14) |
| Right Ventricle SurfaceVolumeRatio | 99.2% | 0.88 (0.78, 1.00) |

##### Supplementary Table 6: Most Stable Radiomic Features for Predicting Myocardial Infarction

| Radiomics Feature | Stability | Adjusted Hazard Ratio (CI) |
|---|---|---|
| Aorta Elongation | 100% | 1.05 (0.94, 1.17) |
| Aorta LeastAxisLength | 100% | 1.16 (1.06, 1.27) |
| Aorta Maximum2DDiameterSlice | 99.5% | 1.19 (1.05, 1.35) |
| Aorta MinorAxisLength | 99.1% | 1.16 (1.02, 1.31) |
| Aorta SurfaceVolumeRatio | 100% | 0.84 (0.71, 0.99) |
| Left Atrium Elongation | 99.7% | 1.13 (1.01, 1.26) |
| Left Atrium Flatness | 100% | 1.33 (1.18, 1.49) |
| Left Atrium Maximum2DDiameterSlice | 98.8% | 1.16 (1.03, 1.32) |
| Left Ventricle LeastAxisLength | 100% | 1.64 (1.45, 1.85) |
| Left Ventricle Maximum2DDiameterSlice | 100% | 1.54 (1.35, 1.77) |
| Left Ventricle Maximum3DDiameter | 91.6% | 1.10 (1.06, 1.14) |
| Left Ventricle MinorAxisLength | 100% | 1.56 (1.43, 1.71) |
| Right Atrium MajorAxisLength | 100% | 1.03 (0.95, 1.12) |
| Right Atrium Maximum2DDiameterColumn | 100% | 1.08 (1.03, 1.14) |
| Right Atrium Sphericity | 100% | 1.27 (1.12, 1.44) |
| Right Atrium SurfaceArea | 99.9% | 1.09 (1.00, 1.20) |
| Right Atrium SurfaceVolumeRatio | 99.7% | 0.71 (0.57, 0.90) |
| Right Ventricle LeastAxisLength | 100% | 1.01 (0.87, 1.17) |
| Right Ventricle Maximum3DDiameter | 100% | 1.06 (1.02, 1.11) |
| Right Ventricle Sphericity | 99.3% | 0.85 (0.75, 9.96) |
| Right Ventricle VoxelVolume | 96.5% | 1.27 (1.11, 1.44) |

**Supplementary Table 7: Most Stable Radiomic Features for Predicting Stroke**

| Radiomics Feature | Stability | Adjusted Hazard Ratio (CI) |
|---|---|---|
| Aorta Elongation | 97.5% | 1.15 (0.93, 1.42) |
| Aorta MinorAxisLength | 100% | 1.35 (1.05, 1.73) |
| Aorta SurfaceVolumeRatio | 100% | 0.56 (0.41, 0.78) |
| Left Atrium Flatness | 100% | 1.35 (1.08, 1.70) |
| Left Atrium LeastAxisLength | 10% | 1.53 (1.20, 1.95) |
| Left Atrium Sphericity | 95.8% | 1.47 (1.11, 1.93) |
| Left Ventricle Maximum2DDiameterSlice | 95.8% | 1.19 (0.89, 1.59) |
| Right Atrium Flatness | 100% | 1.09 (0.88, 1.34) |
| Right Ventricle Flatness | 100% | 0.78 (0.62, 0.99) |
| Right Ventricle MeshVolume | 100% | 1.18 (0.88, 1.58) |
| Right Ventricle Sphericity | 100% | 0.84 (0.65, 1.07) |
| Right Ventricle VoxelVolume | 100% | 1.18 (0.88, 1.58) |

**Supplementary Table 8 : Observed Incident MACE Rates per 1000 Person-Years by Ordinal Radiomics Risk Groups (Rows) and PREVENT Risk Groups (Columns)**

| Radiomics Risk Score | PREVENT Risk Score | | | | |
|---|---|---|---|---|---|
| | **Low** | **Borderline** | **Intermediate** | **High** | **Unknown** |
| **Low** | 4.5 (1.6, 7.8) | 7.8 (1.7, 15.5) | 13.3 (2.8, 25.9) | 0.0 (0.0, 0.0) | 15.0 (12.4, 18.0) |
| **Borderline + Intermediate** | 10.7 (7.4, 17.8) | 11.3 (4.4, 14.4) | 19.9 (9.1, 22.6) | 0.0 (0.0, 0.0) | 19.8 (15.1, 21.5) |
| **High** | 92.8 (46.4, 140.7) | 48.6 (0.0, 90.9) | 39.4 (9.2, 71.0) | 0.0 (0.0, 0.0) | 43.6 (27.6, 61.9) |

**Supplementary Table 9: Observed Incident MACE Rates per 1000 Person-Years by Statin Eligibility Groups Defined by the Radiomics Risk Score (Rows) and the PREVENT Risk Score (Columns)**

| Statin Eligibility by Radiomics Risk Score | Statin Eligibility by PREVENT Risk Score | | |
|---|---|---|---|
| | **Risk < 7.5%** | **Risk ≥ 7.5%** | **PREVENT Missing** |
| **Risk < 5.0%** | 7.3 (5.3, 9.5) | 15.1 (8.7, 21.8) | 16.3 (14.1, 18.6) |
| **Risk ≥ 5.0%** | 28.4 (18.0, 40.7) | 32.1 (19.1, 45.7) | 30.8 (23.5, 38.5) |

#### Supplementary Table 10: NLST Baseline Model Variables

| Feature | Coefficient |
|---|---|
| Age | 0.093 |
| Sex | 0.530 |
| Race | -0.012 |
| Pack-years | 0.005 |
| Current smoking status | 0.745 |
| BMI | 0.019 |
| History of Diabetes | 0.773 |
| History of Hypertension | 0.490 |
| History of Cancer | -0.274 |
